# Implementing AI-assisted, patient-friendly imaging report summaries to enhance oncology care delivery: protocol for a randomized mixed-methods quality improvement study

**DOI:** 10.64898/2026.10.05.26364529

**Authors:** Mary Ryan Baumann, Hanna J. Barton, Paula Voorheis, Corey Dahl, Lori DuBenske, Winifred Commers, Kim Mast, Toby Campbell, Brian W. Patterson, Hamid Emamekhoo

## Abstract

**Background:** As required by the 21st Century Cures Act, all test results, including final radiology reports, are released to patients as soon as the results are finalized. However, radiology reports are primarily intended for communication between medical professionals and can often be difficult for patients to interpret, particularly in cancer care settings. We developed a tool that leverages a large language model (LLM; ChatGPT-5.2, OpenAI, San Francisco, CA) to draft a patient-friendly plain language version of imaging results (Patient-Friendly Scan Result Generation [PFSG] tool) that oncologists can incorporate into their communication with patients via patient portals. This article describes our mixed-methods study protocol.

**Objective:** Our primary aims are to: 1) assess whether patients who receive patient-friendly versions of scan results report better understanding of their results than patients who do not; 2) assess whether introduction of the PFSG tool affects provider workload; and 3) explain how and under what circumstances the PFSG tool enhances communication and workflows, and affects patient and provider outcomes.

**Methods:** We will use an embedded mixed-methods, prospective cohort design within a quality improvement (QI) initiative. The PFSG tool will be implemented using a modified three-wave staircase trial design, with providers in randomized cohorts of oncology disease groups progressively gaining access to the tool over the course of the study. Approximately 60 providers from 11 oncology disease focus groups will be randomized. Longitudinal provider outcomes will be measured through surveys at one month pre-implementation, one month post-implementation, and three months post-implementation. Patient outcomes will be measured cross-sectionally through surveys at one month pre-implementation (n=350) and at post-implementation result point-of-contact (n=350). Additionally, we will conduct interviews with patients and oncologists to inform the iterative design of the tool and its implementation.

**Results:** This study was funded in September 2025 and data collection began in February 2026. As of September 2026, the PFSG tool has been rolled out to all oncology providers, with Cohort 3 post-implementation data collection beginning in August 2026. A total of 738 patient baseline surveys have been collected and patient post-implementation data collection is in progress.

**Conclusions:** This is the protocol for a study of the implementation of a patient-friendly results tool in oncology provider workflows to facilitate improved communication of radiology results to oncology patients. The results of this study will lend evidence to the utility and acceptability of the study tool, and tools like it, to close an important oncology care gap. If the tool is shown to improve key oncology patient and provider outcomes, it may lead to development of a wider release for more general patient populations.

**Trial Registration:** https://clinicaltrials.gov/study/NCT07532083

## INTRODUCTION

As required by the 21st Century Cures Act,^1–3^ all test results, including final radiology reports, are released to patients as soon as the results are finalized. However, these radiology reports are primarily intended for communication between medical professionals and can often be difficult for patients to interpret.^4,5^ A key patient subpopulation affected by this policy is cancer patients, as they frequently undergo radiology imaging studies to assess their disease and treatment response. Due to availability and scheduling limitations, it is common for these scans to be conducted several days before the scheduled follow-up appointment with their oncology provider. In the meantime, reviewing complex radiology reports without the guidance of their healthcare provider can lead to patient confusion and anxiety,^5–9^ resulting in increased queries directed to providers or other healthcare staff.^4,8,10^

Currently, there is no standard recommended workflow for communicating imaging results to cancer patients before their clinic visits. Some providers may send a message to the patient through the electronic health record (EHR) system – often when the results are anticipated – or call them if an unexpected finding is noted or urgent intervention is necessary.^4^ Other providers may simply postpone reviewing the findings to the scheduled visit unless the patient asks about them beforehand. The significance of imaging findings is rarely clear from a single exam alone; interpreting them requires knowledge of the patient’s overall disease course and treatment history, which makes the treating oncologist best positioned to place the results in clinical context and discuss them with the patient, either synchronously or asynchronously.^6^ Patients commonly reach out to the clinical team for clarification, requesting callbacks or sending questions through patient portals;^4^ handling these inquiries and phone calls may add to an oncology provider’s workload. Thus, a solution is needed that should ideally increase patient understanding of their results while not increasing their anxiety or add to clinician burden.

One such solution may be to use artificial intelligence (AI) and large language models (LLMs) to draft lay translations of results to include in patient-facing communications. A 2024 randomized trial by Berigan and colleagues in a general primary care patient population found that providing patients with LLM-generated summaries (Google Gemini, Mountainview, CA) and patient-friendly radiology report translations (Scanslated, Inc, Tampa, FL) significantly improved patient understanding of their imaging reports compared to standard of care.^11^ In addition, a 2026 study that asked participants from a general patient population to view a fixed set of original radiology or AI-generated plain language reports (ChatGPT-4.0, OpenAI, San Francisco, CA) in a randomized sequence found that participants demonstrated higher comprehension accuracy, higher accuracy in follow-up actions, and lower perceived anxiety after viewing the AI-generated plain language reports.^12^ While these efforts demonstrate that AI-generated report translations or summaries are promising in general, to our knowledge, none have investigated solutions specifically tailored to oncology patient care contexts. Further, none investigate how incorporation of such solutions may impact provider workflows.

To address these gaps, we developed a tool that leverages an LLM (ChatGPT-5.2, OpenAI, San Francisco, CA) to draft a patient-friendly plain language version of imaging results that oncologists can incorporate into their communication with patients via patient portals. While its functionality and accuracy were previously piloted in a sandbox environment with a limited number of oncology providers, its utility and acceptability among a wider oncology provider pool or among oncology patients has not yet been assessed. To answer these questions, we designed an embedded randomized, mixed-methods prospective cohort quality improvement (QI) study to measure the tool’s initial impact as it is introduced into an academic learning healthcare system. This article describes our study protocol, including objectives and design considerations.

## METHODS

### Objectives

The broad objectives of this study are three-fold, which we detail below and summarize in **Table 1**.

**Table 1:** Summary of study objectives

| **Table 1: Summary of study objectives** |  |  |  |
| --- | --- | --- | --- |
|  | **Designation** | **Objective** | **Associated Data Collection Methods** |
| Patient Objective | Co-primary | The primary patient-focused aim is to assess whether patients who receive patient-friendly version of scan results report better understanding of their results than patients who do not. | • Pre- & post-implementation surveys of patients |
| Provider Objective | Co-primary | The primary provider-focused aim is to assess whether introduction of the PFSG tool affects provider workload. | • Pre- & post-implementation surveys of physicians |
| Mixed-Methods Objective | Secondary | The mixed-methods aim is to explain how and under what circumstances the PFSG tool enhances communication and workflows, and affects patient and provider outcomes | • Patient interviews<br>• Open-ended patient survey questions<br>• Academic detailing with providers |

First, this study aims to evaluate patient experiences and preferences related to accessing radiology scan results and communicating with providers about those results, including use of MyChart (©Epic Systems, Inc., Verona, WI) and perceptions of AI-assisted, patient-friendly versions of scan results. The primary patient-focused aim is to assess whether patients who receive patient-friendly versions of results report better understanding of their results than patients who do not.

Second, this study aims to evaluate provider experiences, preferences, and feedback related to radiology-result communication workflows and the implementation of a tool that generates patient-friendly versions of scan results (Patient-Friendly Scan Result Generation [PFSG] tool), including perceived usability, workflow impact, and satisfaction. The primary provider-focused aim is to assess whether introduction of the PFSG tool affects provider workload.

Third, this study aims to integrate quantitative patient and provider outcomes with qualitative accounts of communication practices, tool use and non-use, workflow conditions, and implementation experiences. The mixed-methods aim is to explain how and under what circumstances the PFSG tool enhances communication and workflows, and affects patient and provider outcomes.

### Patient-Friendly Scan Result Generation (PFSG) Tool

The tool under investigation is one that allows providers to generate AI-assisted drafts of patient-friendly versions of scan results (PFSG); providers can then edit these drafts, incorporate them into their patient communications, and attach them to patient scan results (see **Figure 1** for a workflow illustration). The PFSG tool will be progressively implemented among 11 oncology disease focus groups (benign hematology, breast, gastrointestinal, general oncology, genitourinary, gynecological, head and neck, lymphoma, melanoma, stem cell transplantation and cellular therapy, thoracic) in a midwestern academic health system over the course of 34 weeks. The PFSG tool uses a secure private instance of OpenAI GenAI models (GPT models) running in the Azure cloud (Microsoft, Redmond, WA). Data passed to the system is not preserved by OpenAI/Azure and is not used for model training in any form. Data transfer and security for this usage was approved by health system security, and AI usage was approved by the hospital’s Clinical AI and Predictive Analytics Committee. The PFSG tool does not access radiology images and only uses the radiology report itself; further, it neither interprets nor analyzes the results and does not provide any clinical recommendations.

**Figure 1:**
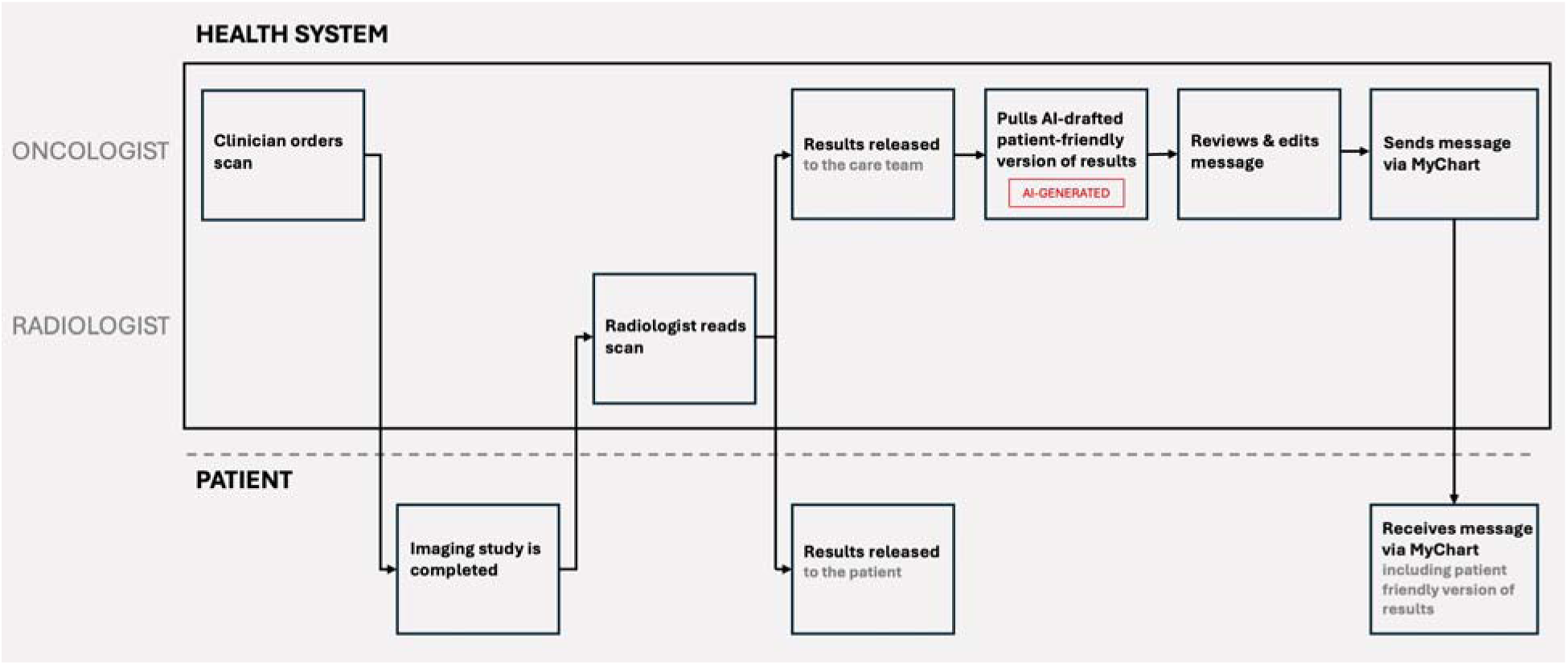
Illustration of Patient-Friendly Scan Result Generation (PFSG) tool workflow from scan order to patient message receipt.

The PFSG tool was previously integrated into routine clinical care workflows in a gated production environment for pilot testing, allowing providers to use it through the Epic standard Results Management workflow. Once providers gain access to the PFSG tool at their randomized implementation timepoint in this study, they will be able to pull an AI-generated patient-friendly version of their patient’s scan results into a MyChart message using a DotPhrase, review the message, and edit as appropriate before sharing with the patient (**Figure 1**).

### Study Setting & Design

This study will be conducted in oncology ambulatory clinics at University of Wisconsin Hospitals and Clinics (UW Health), which serves more than 33,000 patients across southwestern Wisconsin and northern Illinois. This study employs an embedded mixed-methods, prospective cohort design within a QI initiative. The PFSG tool will be implemented using a modified three-wave stepped-wedge or staircase trial design,^13^ with providers in randomized cohorts of oncology disease focus groups progressively gaining access to the PFSG tool over the course of the study (**Figure 2**). Provider outcomes will be measured longitudinally through surveys at three timepoints: one month pre-implementation, one month post-implementation, and three months post-implementation. Patient outcomes will be measured cross-sectionally through surveys at two timepoints: one month pre-implementation and at post-implementation result point-of-contact. Additionally, we will conduct interviews with patients and oncologists to inform the iterative design of the tool and its implementation. This trial has been registered on ClinicalTrials.Gov (NCT07532083). Data collection began in February 2026, and the study is expected to complete data collection by November 2026.

**Figure 2:**
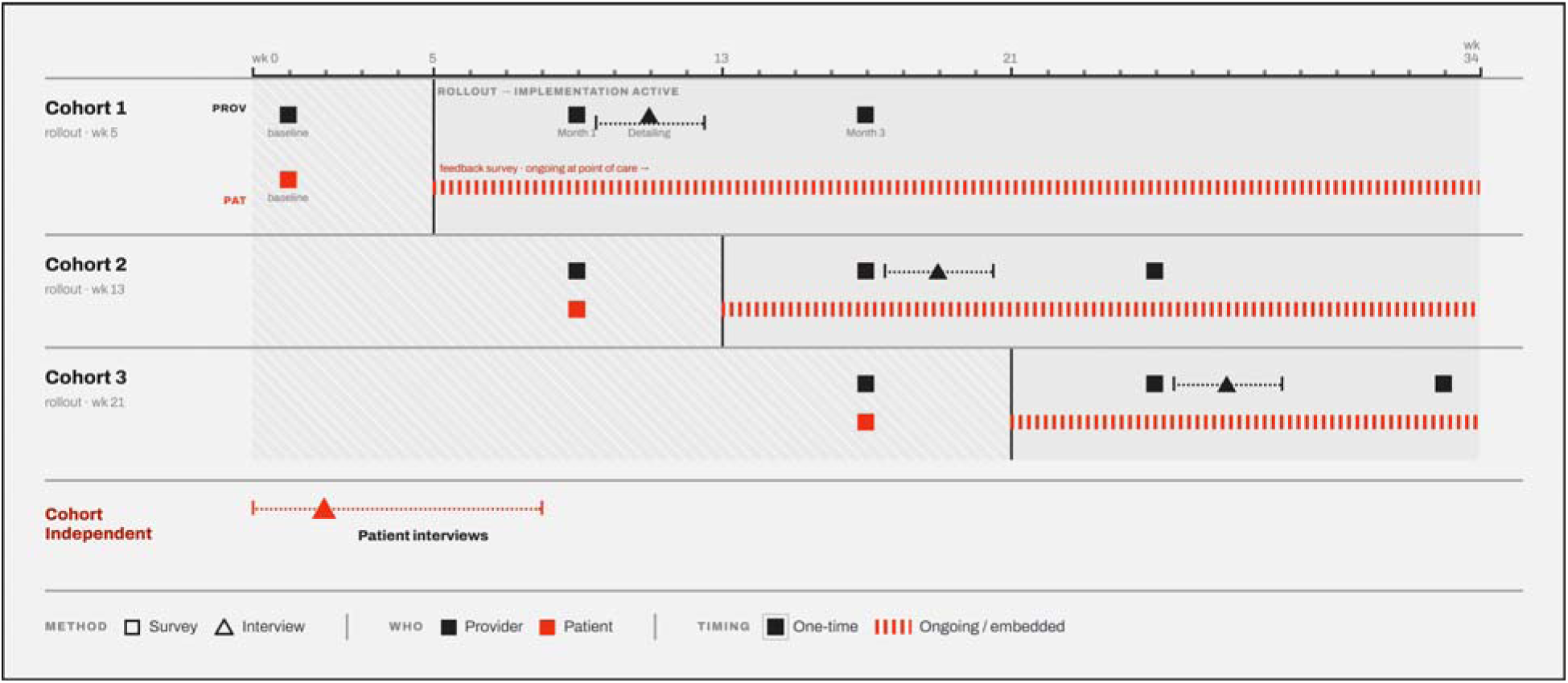
Study design schematic illustrating patient- and provider-level data collection, interviews, and rollout timing across the 34 study weeks. Timelines are stratified by implementation cohort, as well as cohort-independent activities. Black coloring indicates study activities aimed at providers; red coloring indicates study activities aimed at patients. Squares indicate survey activities, while triangles indicate interview activities. Solid points indicate study activities administered at a single timepoint, while dotted or dashed lines indicate embedded and ongoing activities.

### Study Participant Recruitment & Selection Criteria

Oncology patients and providers will be eligible for participation in different components of this study.

Patients who are aged 18 years or older; have a current or prior cancer diagnosis of any cancer type; have active access to the UW Health MyChart patient portal; are receiving care at UW Health within the Division of Hematology, Medical Oncology and Palliative Care or Department of Gynecologic Oncology; are undergoing imaging studies (e.g., CT, MRI, or PET scans) as part of routine cancer care; and are willing and able to complete electronic surveys will be eligible to participant in the patient component of this study. For interview participation, patient participants must also be willing to participate in a 30–45-minute phone or video interview. Patient participants who are unable to provide informed consent due to cognitive impairment or other limitations (as determined by the clinical or study team), or are enrolled in an active interventional clinical trial (except for biospecimen, registry, or non-interventional studies) will be excluded. Patients with radiographic disease progression will be excluded from interview consideration, as to avoid increasing cognitive burden by adding a research interview to their tasks at a time when they will need to make important decisions about treatment modifications with their treating provider. All eligible patients identified in the EHR will receive an invitation to complete a baseline survey, while a subset will be recruited for interview participation. Eligible patients who receive a PFSG summary from their provider will be invited to complete a post-implementation survey via a link embedded in their scan summary MyChart message.

Providers who are faculty physicians (MD or DO) practicing in the Division of Hematology, Medical Oncology and Palliative Care or Department of Gynecologic Oncology at UW Health; are involved in reviewing, discussing, or sharing imaging results with patients; were scheduled to gain access to the PFSG tool as part of eventual general clinical implementation; and are willing and able to complete electronic surveys or participate in a brief 30-minute interview will be eligible to participant in the oncology provider portion of this study. All eligible providers will be invited to participate via departmental emails; clinical study team members will also introduce the project during divisional and group meetings to promote participation. A subset of providers will also be recruited to participate in academic detailing interviews.

### Data Collection

Key patient and provider outcomes are summarized in **Table 2**.

**Table 2:**
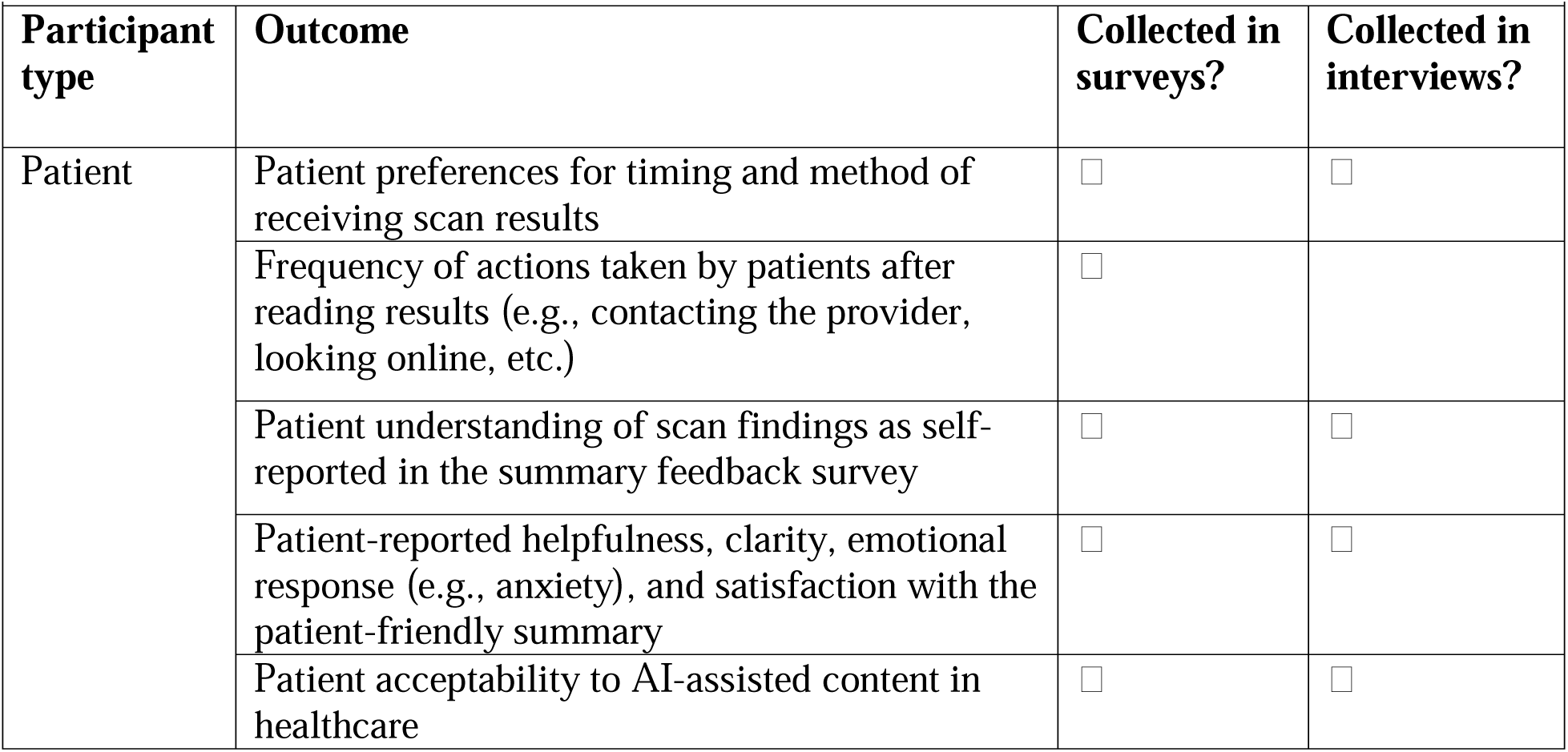
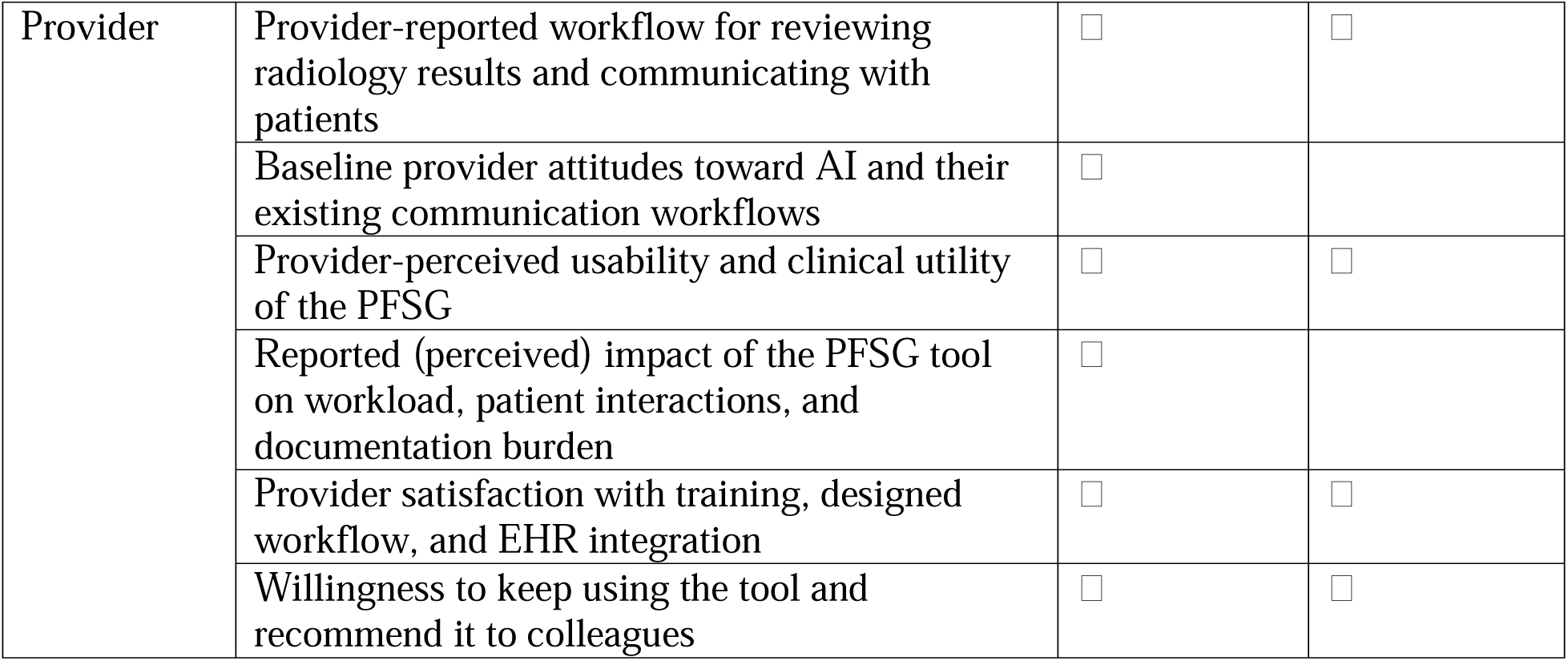
Key patient and provider outcomes collected.

| **Table 2: Key patient and provider outcomes collected.** |  |  |  |
| --- | --- | --- | --- |
| **Participant type** | **Outcome** | **Collected in surveys?** | **Collected in interviews?** |
| Patient | Patient preferences for timing and method of receiving scan results | ☐ | ☐ |
|  | Frequency of actions taken by patients after reading results (e.g., contacting the provider, looking online, etc.) | ☐ |  |
|  | Patient understanding of scan findings as self-reported in the summary feedback survey | ☐ | ☐ |
|  | Patient-reported helpfulness, clarity, emotional response (e.g., anxiety), and satisfaction with the patient-friendly summary | ☐ | ☐ |
|  | Patient acceptability to AI-assisted content in healthcare | ☐ | ☐ |
| Provider | Provider-reported workflow for reviewing radiology results and communicating with patients | ☐ | ☐ |
|  | Baseline provider attitudes toward AI and their existing communication workflows | ☐ |  |
|  | Provider-perceived usability and clinical utility of the PFSG | ☐ | ☐ |
|  | Reported (perceived) impact of the PFSG tool on workload, patient interactions, and documentation burden | ☐ |  |
|  | Provider satisfaction with training, designed workflow, and EHR integration | ☐ | ☐ |
|  | Willingness to keep using the tool and recommend it to colleagues | ☐ | ☐ |

### Patient Data Collection

Patient data will be collected through surveys administered at baseline/pre-implementation and post-implementation point-of-care using REDCap (Research Electronic Data Capture;^14,15^ Multimedia Appendix 1). After screening questions and informed consent, baseline surveys will begin with demographic characteristics before proceeding to sections about previous experiences viewing scan results in MyChart, experiences around communications from their cancer provider regarding scan results, and attitudes toward AI in healthcare. Questions about MyChart scan result experiences include time gaps between scans and follow-up appointments, result viewing habits, confidence in result understanding, and habits for resolving questions about results.

Questions about communication involve contact frequency, communication satisfaction, and communication preferences. Post-implementation surveys will include the same screening and informed consent questions as the baseline survey, with the addition of a question confirming that patients have reviewed the patient-friendly translation generated by the PFSG tool. After questions regarding demographic characteristics, the post-implementation survey will include sections on habits around viewing scan results in MyChart and communicating with their cancer provider regarding scan results, their experience with the patient-friendly summary included in their current scan result, and attitudes toward AI in healthcare. In both surveys, patients have the option to provide an email address for the chance to receive a $20 e-gift card.

Prior to implementation, patients will also be recruited for interviews. We will review upcoming clinic schedules to identify eligible patients to approach for study consent and interview scheduling. Interviews will be conducted via video conferencing software (Zoom, San Jose, CA) and cover patients’ prior experiences with receiving scan results via MyChart. Additionally, we will present the patient with a mock patient scan result translation during the interview and prompt for specific feedback. The interview guide was developed by the interdisciplinary study team which included expertise in oncology, human factors engineering, and qualitative methods, and was informed by critical incident interviewing techniques^15,16^ and the Systems Engineering Initiative for Patient Safety (SEIPS) model^16,17^ (see Multimedia Appendix 2). With permission, interviews will be audio-recorded; otherwise, detailed structured notes will be taken.

### Provider Data Collection

Data will be collected from providers through a survey administered at pre-implementation and two surveys administered post-implementation using REDCap (see Multimedia Appendix 1). Baseline surveys will include questions regarding time gaps between patient scans and follow-up appointments, provider habits in initiating result communication, experiences with patient-initiated communication (including impact on workload), preferences for features in a hypothetical result summary/translation tool, and attitudes toward AI in healthcare. Post-implementation surveys will include similar questions regarding provider habits in initiating result communication and experiences with patient-initiated communication (including impact on workload) based on a one-month recall period; they will also include questions about frequency of PFSG tool use, experiences with using the PFSG tool, and attitudes towards AI in healthcare.

Following PFSG implementation, providers will be invited to participate in individual, semi-structured academic-detailing interviews lasting approximately 20–30 minutes. Using a standardized, theory-informed guide, interviews will examine existing scan-result communication practices; PFSG use and non-use; workflow fit; editing and adaptation of generated summaries; perceived benefits, challenges, accuracy, and trust; effects on patient communication; and recommended improvements. The guide is informed by the Easy, Attractive, Social and Timely (EAST),^18^ SEIPS, and selected Exploration, Preparation, Implementation, Sustainment (EPIS)^19,20^ constructs. With permission, interviews will be audio-recorded; otherwise, detailed structured notes will be taken.

### Randomization & Blinding

Prior to the start of data collection, 11 oncology disease focus groups were randomized as clusters of providers to one of three times when they will receive access to the PFGS tool; the implementation time points are spaced over 8-week intervals after an initial global 4-week standard of care period. Randomization was conducted using a 1:1:1 allocation ratio and covariate constrained randomization to balance by disease group-level scan order volume, disease-group provider count, and anticipated tool utilization; therefore, the number of disease groups per cohort and the number of providers per disease group varies. The randomization scheme was generated by the study statistician using R (v4.4.3)^21^ and implemented in REDCap.

Given the nature of the QI, blinding will not be feasible. Providers will be aware of when they receive access to the PFSG tool as part of their clinical workflow. Patients will not be explicitly told if they will be receiving PFSG summaries with their scan results, but it will be evident when the PFSG tool has been implemented for their provider based on the scan results the patient receives.

### Sample Size & Power

Patient-level sample size was calculated using the primary patient endpoint, self-reported overall understanding of scan results measured via a five-point Likert scale (Very easy; Somewhat easy; Neither easy nor difficult; Somewhat difficult; Very difficult). Assuming 40% of patients at the baseline survey report that the information presented in their scan results are either “Very easy” or “Somewhat easy” to understand, a sample size of 350 patient responses to each survey (baseline and post-implementation) will be able to detect a 12% absolute increase at the post-implementation survey (increase to 52%) with 83% power and a 2.5% two-sided type I error rate using a two-sample t-test.

Provider-level sample size is constrained by the number of oncology providers practicing at UW Health (approximately 60); power was calculated using the primary provider endpoint, perceived impact of scan result patient queries on provider workload measured via a five-point scale (No impact; Minimal impact; Moderate impact; Significant impact; Overwhelming impact). We define clusters as oncology disease focus groups, with individual providers nested in disease groups. We assume a within-provider intracluster correlation of 0.65, a within-period disease-group (cluster) intracluster correlation of 0.03, and a disease-group autocorrection of 0.8. At initial sample size estimation, it was assumed that randomization would be balanced by disease group-level scan order volume, resulting in variation in the number of disease groups per cohort and the number of providers per disease group; for power estimation, we assume 4 disease groups per implementation cohort, with 4 providers per disease group (N=48). We assume that 35% of providers report in the baseline survey that the effect of scan-result-related queries has at most a minimal impact on their clinical workload. Given these assumptions and a 2.5% two-sided type I error rate, we will have 80% power to detect a 24% absolute increase in providers reporting that scan-result-related queries has at most a minimal impact on their clinical workload (increase to 59%); with 5 providers per disease group (N=60), we will have 88% power to detect the same effect size, or 80% to detect an increase to 57% (22% absolute increase).

### Statistical Analysis

We will generate descriptive statistics for all patient and provider participants. Variables collected during the baseline or pre-implementation periods will be considered as baseline variables. Continuous variables will be summarized using the non-missing sample size, the number of missing observations, and mean (standard deviation) or median (interquartile range) as appropriate. The frequency and percentages (based on the non-missing sample size) will be reported for all categorical measures. In general, data will be sorted by implementation cohort. For provider-level data, they will also be sorted by collection timepoint within cohort, when appropriate.

The full analysis patient population will include all invited patients who completed baseline or post-implementation patient surveys, regardless of whether their provider fully adhered to the study protocol; however, we cannot observe post-implementation survey outcomes from patients whose providers did not utilize the PFSG tool, as survey links are only attached at the bottom of result summary reports generated by the PFSG tool. Survey responses from this population will be analyzed using (generalized) linear mixed effects models (GLMMs) adjusted for secular trend via categorical time-period fixed effects, with random intercepts for oncology disease group.

GLMMs will also adjust for baseline disease group-level scan order volume and disease group size to account for variables used in constrained randomization.^22^ Patients are nested within disease group; however, patient data is collected cross-sectionally, so time-specific measurements are not nested within patient. An identity link will be used for continuous outcomes, while a logit link will be used for binary responses. Likert endpoints (1-5 scale) will be dichotomized to “achieves 4-5 rating” or “achieves 1-2 rating”, depending on outcome sentiment (positive/negative). Categorical outcomes will be analyzed using a two-way (pre- vs post-implementation) table Chi-squared test as an exploratory/sensitivity analysis.

The primary provider population will be analyzed according to intention-to-treat principles using GLMMs adjusted for secular trend via categorical time-period fixed effects, with random intercepts for disease group- and provider-by-time period interactions. Similar to the patient-level analyses, GLMMs will also adjust for baseline disease group-level scan order volume and disease group size to account for variables used in constrained randomization. Time-specific measurements will be nested within provider; providers are nested within disease group. An identity link will be used for continuous outcomes, while a logit link will be used for binary responses. Likert endpoints (1-5 scale) will be dichotomized similar to patient Likert endpoints.

Informal patient-level subgroup domains identified for exploration include gender, age, highest educational attainment, preferred language, and implementation cohort. No *a priori* power calculations were performed for detecting specific subgroup differences, so subgroup analyses will be considered exploratory. Subgroup analyses will be conducted by including a main effect term for the subgroup variable, as well as an interaction between study condition and subgroup (treatment subgroup). Subgroup-specific treatment effects will be estimated via stratified analyses and visualized using forest plots.

To control for false discovery rate between the patient- and provider-focused arms, we will use a two-sided type I error rate of 2.5% for the co-primary hypotheses. Exploratory hypotheses will use a 5% type I error threshold for significance with no correction for multiple testing.

All statistical analyses will be performed using R Version 4.4.3 or higher on a MacOS platform.^21^

### Qualitative Analysis

Patient interviews will be analyzed in two segments corresponding to two domains addressed in the interview guide. First, interview questions pertaining to the PFSG prototype will be extracted and analyzed using rapid qualitative analysis,^23–25^ allowing for timely synthesis of patient perspectives to inform iterative tool development. Second, sections of the interview addressing patients’ prior experience receiving scan results through MyChart will be analyzed using a combined inductive and deductive approach, guided by SEIPS 3.0,^16,17^ selected for its focus on the longitudinal, or “patient journey,” aspect of care. This hybrid content analysis^26,27^ will identify system factors (e.g., person, tasks, tools and technology, organization, environment) that shaped patients’ prior experiences accessing scan results, developing our understanding of how scan result communication fits into the broader sociotechnical context of their cancer care journey.

Provider academic detailing interviews will be analyzed using rapid qualitative analysis adapted from Hamilton and colleagues.^23,24^ Data will be summarized using a structured template aligned with the interview guide and the EAST, SEIPS, and EPIS frameworks to identify barriers and facilitators influencing providers’ use of the PFSG tool, including individual, workflow, organizational, and implementation factors. Findings will then be transferred to a participant-by-domain matrix. Analysts will pilot the template on initial interviews; they will also compare findings within and across providers to distinguish barriers and facilitators that are similar versus differing, locate contextual factors explaining these differences, and identify actionable opportunities to improve PFSG implementation and use.

### Mixed Methods Analysis

Quantitative and qualitative data will be analyzed separately at their designated points in the intervention and integrated according to the purpose of each qualitative component. Patient interviews will enhance the intervention by informing iterative refinement of the PFSG prototype and identifying patient-level and sociotechnical factors shaping how scan results are accessed and experienced across the cancer care journey. Provider academic-detailing interviews will enhance the experimental findings by explaining tool use, workflow experiences, and quantitative outcomes, while also informing improvements to training and implementation support. Integrated conclusions will describe how the qualitative findings enhanced both the intervention and interpretation of the experimental result.

## RESULTS

This study was funded in September 2025 and ethics approval was obtained in December 2025 (University of Wisconsin Institutional Review Board #UW25122). The trial was registered on ClinicalTrials.gov in April 2026. Data collection began in February 2026. As of September 2026, the PFSG tool has been opened to all oncology providers, with Cohort 3 post-implementation data collection beginning in August 2026. A total of 738 patient baseline surveys have been collected; patient post-implementation data collection is in progress with 31 completed as of September 2026. Data collection is anticipated to be completed by November 2026.

## DISCUSSION

This article outlines a study protocol for the implementation of a patient-friendly result translation tool in oncology provider workflows to facilitate improved communication of radiology results to oncology patients. We hypothesize that the implementation of the PFSG tool will result in increased self-reported oncology patient understanding of their radiology results while not substantially increasing oncology provider burden. While previous efforts have examined the impact of AI-and LLM-based tools in general patient populations, none have specifically investigated oncology care contexts where patients are often highly engaged,^28^ their results can be nuanced and complex depending on the patient’s treatment course and disease history, and workflows are unique from other specialties. Further, while heavy clinician workloads and burnout is a widespread issue across healthcare specialties,^29^ it is particularly evident among oncologists.^30,31^

Other technologies similar to the PFSG tool are beginning to become available. For example, Epic has recently developed a built-in AI assistant and chatbot on the patient-facing side (Emmie) that would allow patients to generate a translation or summary of clinician-provided results or communication and answer questions without the need to add to provider workload.^12,32^ However, this represents a “black box” from the provider side, as providers have no insight into what information the assistant is providing (or expectation of what information it would provide) and no way to control contextualization of translations. By incorporating a human-in-the-loop approach, the PFSG tool under investigation in this study allows for provider agency while supplying easier-to-understand lay translations of results to patients.

We note several limitations of this study. First, while we will be able to collect longitudinal data from oncology providers, we have limited ability to obtain longitudinal information from oncology patients. As the study is embedded within a QI initiative, it was difficult to determine at study-onset which patients with previous experience of radiology scan result communications would also be likely to receive radiology scan results during the post-implementation periods; thus, patient data is not linked between collection timepoints. Second, as this study is designed to be embedded within a QI initiative and adhere to actual practice workflows as much as possible, use of the PFSG tool is not mandatory for any provider and is instead offered as a possible communication tool at the provider’s disposal. This means it is possible that there may be low tool uptake or utilization that may limit post-implementation data collection opportunities. The study team is proactively combatting this possibility by holding pre-implementation “onboarding” information sessions for upcoming disease groups to increase awareness of the tool’s availability and answer questions; further, academic detailing sessions allow the study team to identify pain points in provider workflows and offer troubleshooting advice.

The results of this study will lend evidence to the utility and acceptability of the PFSG tool, and tools like it, to close an important oncology care gap. Further, academic detailing and interview results will allow for further refinement of the tool. If the tool is shown to improve key oncology patient and provider outcomes, it may lead to development of a wider release for more general patient populations.

## Supporting information

Multimedia Appendix 2

Multimedia Appendix 1

Multimedia Appendix 3

## Data Availability

Deidentified versions of the survey datasets generated and analyzed during this study will be available upon request after primary analyses have been completed and primary results have been published. Patient and clinician interview data will not be made available due to privacy concerns.

## ACKNOWLEDGEMENTS

The authors used Anthropic Claude Design to generate and revise figures. No other AI tools were used for content generation or editing. The authors reviewed and edited the output as needed and take full responsibility for the content of the publication.

## FUNDING

This work was supported by funding from the University of Wisconsin Hospital and Clinics and the National Institutes of Health Clinical and Translational Science Award (NIH/NCATS UL1TR002737).

## CONFLICTS OF INTEREST

WC reports spousal employment at Epic Systems. The other authors declare no relevant conflicts of interest.

## AUTHOR CONTRIBITIONS

Conceptualization: HE

Data curation: HJB, MRB, PV, WC

Formal analysis: HJB, MRB, PV

Funding acquisition: HE

Investigation: BWP, HE, HJB, PV, WC

Methodology: HE, HJB, LD, MRB, PV

Project administration: CD

Supervision: BWP, HE, KM, TC

Visualization: HJB, MRB

Writing – original draft: MRB, HJB, PV

Writing – review & editing: BWP, CD, HE, HJB, KM, LD, MRB, PV, TC, WC

## ABBREVIATIONS

**AI** Artificial intelligence

**CT** Computed tomography

**DO** Doctor of osteopathic medicine

**EAST** Easy, Attractive, Social and Timely

**EHR** Electronic health record

**EPIS** Exploration, Preparation, Implementation, Sustainment

**GLMM** Generalized linear mixed model

**LLM** Large language model

**MD** Doctor of medicine

**MRI** Magnetic resonance imaging

**PET** Positron emission tomography

**PFSG** Patient-Friendly Scan Result Generation

**QI** Quality improvement

**REDCap** Research Electronic Data Capture

**SEIPS** Systems Engineering Initiative for Patient Safety

**UW Health** University of Wisconsin Hospitals and Clinics

## MULTIMEDIA APPENIX 1

Patient and provider REDCap survey templates.

## MULTIMEDIA APPENDIX 2

Interview guide.

## MULTIMEDIA APPENDIX 3

SPIRIT-AI checklist.

## Notes

### Clinical Trial

NCT07532083

### Author Declarations

The University of Wisconsin Institutional Review Board (#UW25122) gave ethical apporval for this work.

