## Supplementary material for "Implementing AI-assisted, patient-friendly imaging report summaries to enhance oncology care delivery: protocol for a randomized mixed-methods quality improvement study": Multimedia Appendix 2

**Supplementary Material: Patient Interview Guide**

**Section 1: Past experiences receiving scan results through MyChart**

1. Now, think about the last time you received scan results through MyChart—can you walk me through what that looked like?
a. What kind of scan was it?
b. When did you get the alert about your results? How did that alert come through (text, email)? Where were you?
   a. How soon after your scan were you receiving your results? When did you get the scan done?
c. Did you view your results immediately upon receiving that alert? What drove you to view them right then? (curiosity, eagerness, fear, anxiety?)
   a. If not, when did you view them? Why did you wait?
d. Did you view them through the mobile app or on a desktop?
e. When you viewed your results, what were your initial reactions?
   a. Could you understand what they were communicating?
   b. What was the emotional experience of reading the results?
   c. Did you have any questions?
      i. If you did, what did you do with them? (write them down for next Onc appt, follow up with doc via MyChart message?)
   d. Was there a message from your doctor with the results?
      i. If so, how helpful or reassuring was it?
f. If you viewed the results before you got the message from your doctor, do you recall how long you had to wait for those? How were you alerted?
   a. When you did receive the message from your doctor, how helpful or reassuring was it?
g. Is there anything else you want me to know about what that process looked and felt like for you?
2. Now I want to talk a little bit about how the last time you accessed scan results in MyChart compared to previous times for you. In what ways was it similar or different from other times you’ve accessed scan results?
a. Were there times where you felt the results included all of the information you were interested in receiving? Or times when they did not?
b. Were there times where you felt more or less relief or reassurance? Or more or less anxiety? What do you think contributed to that?
c. Were there any times where you felt especially driven to communicate with your provider? Or times where that wasn’t necessary? Why was that?
d. Were there times where you did not receive the doctor’s note? Or times where the doctor’s note was less useful or reassuring? Why was that?
e. Do you typically view results on the mobile app or on a desktop? Overall, were there times where you were more or less satisfied with the process of receiving scan results?

Additional probes:

- Did it matter where you were?
- Did it matter who you were with?
- Was the overall good or bad nature of the results a major factor in your ability to comprehend or be satisfied with the communication?

3. If you were to suggest ways to improve the patient experience of receiving these results, what would you recommend?
a. How could the technology work better?
b. What could the doctor do better?
c. What would you suggest to other patients?
4. Is there anything else you’d like to share about your experiences with scan results through MyChart?

**Section 2: Feedback on a new design**

*Researcher screen-shares a mocked up patient friendly scan version.*

5What are your initial thoughts about the new design?

- Does it include the information that you expect or want to see?
- What are your thoughts about the content of the first paragraph?
- What are your thoughts about the content of the second paragraph?
- What are your thoughts about the content of the third paragraph? Do you think this design would increase / reduce any anxiety associated with getting these results through MyChart? Or provide some reassurance? Why is that?
- Do you think you’d be more, less, or equally as likely to contact your provider via MyChart to follow up? Why would that be?
- Would you say, if you received this message, you’d be more, less, or just as satisfied with the communication? Why is that?
- Do you think this is an appropriate use of artificial intelligence (AI)?

Do you have any other feedback on the content of the AI-generated scan summary?

**Section 3: Demographics**

1. What is your gender identity?
2. What is your age?
3. What is your race?
4. Are you of Hispanic, Latino, or Spanish origin?
5. What is the highest level of education you have completed?
6. Is English your native language?
7. What language do you prefer to use when discussing your healthcare?
8. Do you have regular access to the internet?
9. What level of knowledge do you have of Artificial Intelligence (AI) in healthcare?

1 being “No Knowledge” to 5 being “Expert-level Knowledge”

10. How comfortable are you with the use of AI in healthcare?

1 being “Very Uncomfortable” to 5 being “Very Comfortable”

11. How would you rate your familiarity with navigating healthcare systems?

1 being “Not at all familiar” to 5 being “Very Familiar”

12. How would you describe your familiarity with receiving and viewing scan results in MyChart?

1 being “Not at all familiar” to 5 being “Very Familiar”
