## Supplementary material for "Implementing AI-assisted, patient-friendly imaging report summaries to enhance oncology care delivery: protocol for a randomized mixed-methods quality improvement study": Multimedia Appendix 1

### Screening questions

To help us determine whether this research study is a good fit for you, we'd like to begin with a few brief questions. Please answer them as honestly and accurately as you can.

Thank you for your time and interest!

| | |
|---|---|
| Do you currently have cancer, or have you had cancer in the past? | ◯ I currently have cancer and am receiving treatment or follow-up care<br>◯ I had cancer in the past, but am now cancer-free<br>◯ I have never had cancer |
| Do you undergo intermittent scans (such as CT scans, PET scans, or MRI) for monitoring of your condition? | ◯ Yes<br>◯ No |
| Have you reviewed your scan results in MyChart at least once in the past year? | ◯ Yes<br>◯ No |
| Have you already completed a survey for this study before? | ◯ Yes<br>◯ No |

### Study Information and Informed Consent

Thank you for your interest in this research study.
Please review the following information to help you decide whether you'd like to take part.

What is this study about?
This study explores how patients like you view and interpret their imaging (scan) results in MyChart. We're also studying whether providing a simple, plain-language summary of your scan results can improve understanding and reduces anxiety before your clinic visit.

What will I be asked to do?

Complete a short survey (about 5-10 minutes).
Answer questions about how and when you view your scan results, how confident you feel in understanding them, and your thoughts on receiving simple summaries.
Some patients may receive a simple summary of their scan results in the future and may be invited to complete an additional survey about that experience.

That's it! No clinic visits, follow-ups, or additional tasks are required.

Your participation is voluntary. You may choose to stop at any time. Deciding not to participate will not affect your medical care or any benefits you receive.

Privacy and confidentiality
Your responses will be confidential and used only by the research team.
We will not share your name or other identifying details.
Once the study is complete, any information that could identify you will be removed.
With appropriate confidentiality protections, we might use information that we collect during this study for other research or share it with other researchers without additional consent from you.
A description of this clinical trial will be available on http://www.ClinicalTrials.gov. This website will not include information that can identify you. At most, the website will include a summary of the results. You can search this website at any time.

Use of Health Information:
This study will only use the health information you choose to share with the research team.
Protected health information (PHI) refers to any health information that includes your name or other identifiers.
Your authorization to use your PHI for this research does not have an end date, but you may withdraw it at any time. To withdraw your authorization, you should inform our research team in writing of your intent to withdraw.
If you choose to withdraw your authorization, you will no longer be able to participate in the study. To do so, simply notify the research team in writing.

Risks and benefits
There are no expected risks from completing this survey.
You won't benefit directly, but your input will help improve how scan results are communicated to the patients in the future.

Questions?

If you have questions, concerns, or complaints, or think that participating in the research has hurt you , please contact our research office via email or phone: 608-263-6002.

Or you may reach out to the Principal Investigator, Dr. Toby Campbell,through the research office.

If you have concerns about your rights as a research participant or have complaints about the research study or study team, call the confidential research compliance line at 1-833-652-2506. UW Staff not part of the study team will work with you to address concerns and assist in resolving any complaints.

Email is generally not a secure way to communicate about your health, as there are many ways for unauthorized users to access email. You should avoid sending sensitive, detailed personal information by email. Email should also not be used to convey information of an urgent nature. If you need to talk to someone immediately, please contact Dr. Toby Campbell at 608-263-6002. You do not have to provide your email address to participate in this study.

| | |
|---|---|
| Now that you have more information about the study, would you like to participate and complete the survey? | ◯ Yes<br>◯ No |

Completing this survey will take approximately 10 to 15 minutes. Is this a suitable time to begin the survey?

### Baseline Survey

Thank you for taking the time to participate in this survey.

#### Let's start with a few questions about you.

Who is completing this survey?

- ◯ I am the patient completing this survey about my own experience.
- ◯ I am a family member or caregiver answering this survey on behalf of the patient (e.g., the patient is unable or prefers not to complete surveys).
- ◯ I am a family member or caregiver who regularly reviews the patient's scan results in MyChart and am answering based on my own experience and perspective.
- ◯ I am a healthcare proxy, legal guardian, or other authorized representative completing this survey.
- ◯ Other

Please tell us who is completing the survey if you selected "Other" above.

__________________________________

What is your current age in years?

__________________________________

What is your current gender identity?

- ◯ Man
- ◯ Woman
- ◯ Transgender man
- ◯ Transgender woman
- ◯ Non-binary/gender queer
- ◯ Other
- ◯ Prefer not to answer

Are you of Hispanic, Latino, or Spanish origin?

- ◯ No, not of Hispanic, Latino, or Spanish origin
- ◯ Yes, Mexican, Mexican American, or Chicano/a
- ◯ Yes, Puerto Rican
- ◯ Yes, Cuban
- ◯ Yes, another Hispanic, Latino, or Spanish origin
- ◯ Prefer not to answer

Please select the racial/ethnic category or categories with which you most closely identify.
(Select all that apply)

- ☐ American Indian or Alaska Native
- ☐ Asian or Asian American
- ☐ Black or African American
- ☐ Hispanic or Latino
- ☐ Native Hawaiian or Pacific Islander
- ☐ Middle Eastern or North African
- ☐ White
- ☐ Prefer not to answer

What is the highest level of education you have completed?

- ◯ Less than high school
- ◯ High school diploma or GED
- ◯ Some college, no degree
- ◯ Associate degree (e.g., AA, AS)
- ◯ Bachelor's degree (e.g., BA, BS)
- ◯ Graduate or professional degree (e.g., MA, MS, PhD, MD, JD)
- ◯ Prefer not to answer

Is English your native language?

- ◯ Yes
- ◯ No
- ◯ Prefer not to answer

What language do you prefer to use when discussing your healthcare?

- ◯ English
- ◯ Spanish
- ◯ Hmong
- ◯ Chinese (Mandarin or Cantonese)
- ◯ Arabic
- ◯ Another language
- ◯ Prefer not to answer

Do you have regular access to the internet?

- ◯ Yes
- ◯ No
- ◯ Prefer not to answer

**The following questions relate to your experience viewing scan results in MyChart, as well as communications with your cancer care provider regarding those results.**

How often do you get your scans done on a different day before your follow-up cancer provider appointment?

- ◯ Almost always (they are always done at least a day in advance)
- ◯ Often
- ◯ Sometimes
- ◯ Rarely
- ◯ Never (they are always done the same day)

Typically, how much time is there between your scan and your provider appointment? Usually I get the scans done ...

- ◯ Within 1 week before my appointment
- ◯ Within 2-3 weeks before my appointment
- ◯ More than 3 weeks before my appointment

When your scan results are available to you in MyChart ahead of your provider appointment, how often do you review the results before your appointment?

- ◯ Almost always
- ◯ Often
- ◯ Sometimes
- ◯ Rarely
- ◯ I never review the scan results before my provider appointment.

How important is it for you to be able to review your scan results before meeting with your provider?

- ◯ Not at all important
- ◯ Slightly important
- ◯ Important
- ◯ Fairly important
- ◯ Extremely important

How do you usually review your scan results when they are released to you in MyChart?

(Select the option that best reflects your usual approach.)

- ◯ I review them right away, as soon as I get the notification.
- ◯ I wait until I'm at home or in a calmer setting to read them.
- ◯ I wait to review them with a family member or friend.
- ◯ I do not usually read the scan reports in MyChart.
- ◯ Other

If you chose "Other," please explain how you review your scan results.

__________________________________

How confident are you in your own ability to understand the medical information in your scan results?

- ◯ Not at all confident
- ◯ A little confident
- ◯ Somewhat confident
- ◯ Quite a bit confident
- ◯ Extremely confident

| | |
|---|---|
| When you review the scan results yourself, how easy or difficult is it to understand the information presented in the results? | ◯ Very easy to understand<br>◯ Somewhat easy to understand<br>◯ Neither easy nor difficult to understand<br>◯ Somewhat difficult to understand<br>◯ Very difficult to understand |
| What do you typically do if you have questions about the scan results?<br><br>(Select all that apply) | ☐ Call my provider with my questions<br>☐ Send MyChart messages to my provider with my questions<br>☐ Search online (for example, Google) to get answers<br>☐ Use Artificial Intelligence (AI) tools like ChatGPT to get answers<br>☐ Reach out to family and friends with my questions<br>☐ Wait to discuss the results at my follow-up appointment with my cancer provider and ask my questions then<br>☐ I don't usually have questions |
| When you review the scan results yourself, how often do you contact your provider (e.g., by phone or MyChart message) to ask questions or get clarification before your follow-up appointment with your cancer provider? | ◯ Almost always<br>◯ Often<br>◯ Sometimes<br>◯ Rarely<br>◯ Never<br>◯ I don't review the scan results before my provider appointment. |
| When you review your scan results before your follow-up appointment with your cancer provider, how does it typically affect your anxiety level? | ◯ I usually feel more anxious after reading the results<br>◯ I usually feel less anxious after reading the results<br>◯ My anxiety level doesn't really change<br>◯ I am not sure, or I don't remember<br>◯ Prefer not to answer |
| How satisfied are you with how your scan results are currently shared and explained to you?<br><br>(This includes receiving the scan result in MyChart and any follow-up communication from your care team, if provided.) | ◯ Very satisfied<br>◯ Somewhat satisfied<br>◯ Neutral / No strong opinion<br>◯ Somewhat dissatisfied<br>◯ Very dissatisfied |
| Please tell us more about what works well or what could be improved about how your scan results are currently shared and explained to you: | __________________________________ |
| What would be your preferred way to receive explanations and communications about your scan results? | ◯ In-person during my follow-up appointment with my cancer provider<br>◯ By phone call from my provider<br>◯ Through MyChart written messages<br>◯ Through a patient-friendly summary (explained in simple terms) alongside the original scan report.<br>◯ I do not want additional information beyond the official scan report |
| If you received a summary of the scan findings written in simpler terms, how helpful would you find it? | ◯ Extremely helpful<br>◯ Very helpful<br>◯ Somewhat helpful<br>◯ Slightly helpful<br>◯ Not helpful at all |

| | |
|---|---|
| What level of knowledge do you have of Artificial Intelligence (AI) in healthcare? | ◯ No knowledge<br>◯ Minimal knowledge<br>◯ Some knowledge<br>◯ A great deal of knowledge<br>◯ Expert-level knowledge |
| How comfortable are you with the use of AI in healthcare? | ◯ Very uncomfortable<br>◯ Uncomfortable<br>◯ Somewhat comfortable<br>◯ Comfortable<br>◯ Very comfortable |
| Percentage of answered high-yield questions. | __________________________ |

### Claim your e-gift card

Please enter a valid email address if you would like to receive a $20 e-gift card. ______________________

### Gift card distribution

| | |
|---|---|
| Gift card was sent to this participant? | ◯ Yes<br>◯ No |
| Date when the gift card was sent. | __________________________ |

### Screening questions

To help us determine whether this research study is a good fit for you, we'd like to begin with a few brief questions. Please answer them as honestly and accurately as you can.

Thank you for your time and interest!

| | |
|---|---|
| Do you currently have cancer, or have you had cancer in the past? | ◯ I currently have cancer and am receiving treatment or follow-up care<br>◯ I had cancer in the past, but am now cancer-free<br>◯ I have never had cancer |
| Do you undergo intermittent scans (such as CT scans, PET scans, or MRI) for monitoring of your condition? | ◯ Yes<br>◯ No |
| Have you reviewed your scan results in MyChart at least once in the past year? | ◯ Yes<br>◯ No |
| Did you review the "simple summary" of the scan result that was shared with you by your provider today? | ◯ Yes<br>◯ No |

Some parts of this survey pertain to your experience with the "simple summary" of the scan results shared with you. Please take a moment to review the summary before beginning the survey.

| | |
|---|---|
| Have you ever provided feedback on one of these simple summaries through this survey? | ◯ Yes<br>◯ No |

We welcome and would appreciate your additional feedback on the scan summary. However, please note that the e-gift card is limited to one per participant.

### Study Information and Informed Consent

Thank you for your interest in this research study.
Please review the following information to help you decide whether you'd like to take part.

What is this study about?
This study explores how patients like you view and interpret their imaging (scan) results in MyChart. We're also studying whether providing a simple, plain-language summary of your scan results can improve understanding and reduces anxiety before your clinic visit.

What will I be asked to do?

Complete a short survey (about 5-10 minutes).
Answer questions about how and when you view your scan results, how confident you feel in understanding them, and your thoughts on receiving simple summaries.
Some patients may receive a simple summary of their scan results in the future and may be invited to complete an additional survey about that experience.

That's it! No clinic visits, follow-ups, or additional tasks are required.

The first 125 participants who complete this survey will be eligible for a $20 e-gift card (optional).

Your participation is voluntary. You may choose to stop at any time. Deciding not to participate will not affect your medical care or any benefits you receive.

Privacy and confidentiality
Your responses will be confidential and used only by the research team.
We will not share your name or other identifying details.
Once the study is complete, any information that could identify you will be removed.
With appropriate confidentiality protections, we might use information that we collect during this study for other research or share it with other researchers without additional consent from you.
A description of this clinical trial will be available on http://www.ClinicalTrials.gov. This website will not include information that can identify you. At most, the website will include a summary of the results. You can search this website at any time.

Use of Health Information:
This study will only use the health information you choose to share with the research team.
Protected health information (PHI) refers to any health information that includes your name or other identifiers.
Your authorization to use your PHI for this research does not have an end date, but you may withdraw it at any time.
If you choose to withdraw your authorization, you will no longer be able to participate in the study. To do so, simply notify the research team in writing.

Risks and benefits
There are no expected risks from completing this survey.
You won't benefit directly, but your input will help improve how scan results are communicated to the patients in the future.

Questions?

If you have questions, concerns, or complaints, or think that participating in the research has hurt you , please contact our research office via email or phone: 608-263-6002.

Or you may reach out to the Principal Investigator, Dr. Toby Campbell, through the research office.

If you have concerns about your rights as a research participant or have complaints about the research study or study team, call the confidential research compliance line at 1-833-652-2506. UW Staff not part of the study team, will work with you to address concerns and assist in resolving any complaints.

Email is generally not a secure way to communicate about your health, as there are many ways for unauthorized users to access email. You should avoid sending sensitive, detailed personal information by email. Email should also not be used to convey information of an urgent nature. If you need to talk to someone immediately, please contact Dr. Toby Campbell at 608-263-6002. You do not have to provide your email address to participate in this study.

Now that you have more information about the study, would you like to participate and complete the survey?

◯ Yes
◯ No

Completing this survey will take approximately 10 to 15 minutes. Is this a suitable time to begin the survey?

### Summary Feedback Survey

Thank you for taking the time to participate in this survey.

#### Let's start with a few questions about you.

Who is completing this survey?

- ◯ I am the patient completing this survey about my own experience.
- ◯ I am a family member or caregiver answering this survey on behalf of the patient (e.g., the patient is unable or prefers not to complete surveys).
- ◯ I am a family member or caregiver who regularly reviews the patient's scan results in MyChart and am answering based on my own experience and perspective.
- ◯ I am a healthcare proxy, legal guardian, or other authorized representative completing this survey.
- ◯ Other

Please tell us who is completing the survey if you selected "Other" above.

__________________________________

What is your current age in years?

__________________________________

What is your current gender identity?

- ◯ Man
- ◯ Woman
- ◯ Transgender man
- ◯ Transgender woman
- ◯ Non-binary/gender queer
- ◯ Other
- ◯ Prefer not to answer

Are you of Hispanic, Latino, or Spanish origin?

- ◯ No, not of Hispanic, Latino, or Spanish origin
- ◯ Yes, Mexican, Mexican American, or Chicano/a
- ◯ Yes, Puerto Rican
- ◯ Yes, Cuban
- ◯ Yes, another Hispanic, Latino, or Spanish origin
- ◯ Prefer not to answer

Please select the racial/ethnic category or categories with which you most closely identify.
(Select all that apply)

- ☐ American Indian or Alaska Native
- ☐ Asian or Asian American
- ☐ Black or African American
- ☐ Hispanic or Latino
- ☐ Native Hawaiian or Pacific Islander
- ☐ Middle Eastern or North African
- ☐ White
- ☐ Prefer not to answer

What is the highest level of education you have completed?

- ◯ Less than high school
- ◯ High school diploma or GED
- ◯ Some college, no degree
- ◯ Associate degree (e.g., AA, AS)
- ◯ Bachelor's degree (e.g., BA, BS)
- ◯ Graduate or professional degree (e.g., MA, MS, PhD, MD, JD)
- ◯ Prefer not to answer

Is English your native language?

- ◯ Yes
- ◯ No
- ◯ Prefer not to answer

| | |
|---|---|
| What language do you prefer to use when discussing your healthcare? | ◯ English<br>◯ Spanish<br>◯ Hmong<br>◯ Chinese (Mandarin or Cantonese)<br>◯ Arabic<br>◯ Another language<br>◯ Prefer not to answer |
| Do you have regular access to the internet? | ◯ Yes<br>◯ No<br>◯ Prefer not to answer |

**Section 1: Your past experience with scan results**

**The following questions ask about how you reviewed scan results you received in the past, not the most recent results that came with the "simple summary."**

| | |
|---|---|
| How often do you get your scans done on a different day before your follow-up cancer provider appointment? | ◯ Almost always (they are always done at least a day in advance)<br>◯ Often<br>◯ Sometimes<br>◯ Rarely<br>◯ Never (they are always done the same day) |
| Typically, how much time is there between your scan and your provider appointment? Usually I get the scans done ... | ◯ Within 1 week before my appointment<br>◯ Within 2-3 weeks before my appointment<br>◯ More than 3 weeks before my appointment |
| When your scan results are available to you in MyChart ahead of your provider appointment, how often do you review the results before your appointment? | ◯ Almost always<br>◯ Often<br>◯ Sometimes<br>◯ Rarely<br>◯ I never review the scan results before my provider appointment. |
| How important is it for you to be able to review your scan results before meeting with your provider? | ◯ Not at all important<br>◯ Slightly important<br>◯ Important<br>◯ Fairly important<br>◯ Extremely important |
| How do you usually review your scan results when they are released to you in MyChart?<br><br>(Select the option that best reflects your usual approach.) | ◯ I review them right away, as soon as I get the notification.<br>◯ I wait until I'm at home or in a calmer setting to read them.<br>◯ I wait to review them with a family member or friend.<br>◯ I do not usually read the scan reports in MyChart.<br>◯ Other |
| If you chose "Other," please explain how you review your scan results. | __________________________________ |
| How confident are you in your own ability to understand the medical information in your scan results? | ◯ Not at all confident<br>◯ A little confident<br>◯ Somewhat confident<br>◯ Quite a bit confident<br>◯ Extremely confident |

| | |
|---|---|
| What do you typically do if you have questions about the scan results?<br><br>(Select all that apply) | ☐ Call my provider with my questions<br>☐ Send MyChart messages to my provider with my questions<br>☐ Search online (for example, Google) to get answers<br>☐ Use Artificial Intelligence (AI) tools like ChatGPT to get answers<br>☐ Reach out to family and friends with my questions<br>☐ Wait to discuss the results at my follow-up appointment with my cancer provider and ask my questions then<br>☐ I don't usually have questions |
| When you review the scan results yourself, how often do you contact your provider (e.g., by phone or MyChart message) to ask questions or get clarification before your follow-up appointment with your cancer provider?<br><br>1. (Select the option that best reflects your past experience.) | ◯ Almost always<br>◯ Often<br>◯ Sometimes<br>◯ Rarely<br>◯ Never<br>◯ I don't review the scan results before my provider appointment. |
| What is your preferred way to receive explanations and communications about your scan results? | ◯ In-person during my follow-up appointment with my cancer provider<br>◯ By phone call from my provider<br>◯ Through MyChart written messages<br>◯ A patient-friendly summary (explained in simple terms) in addition to the original scan report.<br>◯ I do not want additional information beyond the official scan report |

**Section 2: Your experience with the "simple summary"**

**The following questions focus on the scan results you just received and the "simple summary" that came with them.**

| | |
|---|---|
| When you reviewed the scan result along with the provided simple summary, how easy or difficult was it to understand the information presented in the scan results? | ◯ Very easy to understand<br>◯ Somewhat easy to understand<br>◯ Neither easy nor difficult to understand<br>◯ Somewhat difficult to understand<br>◯ Very difficult to understand |
| How satisfied are you with how this scan result was shared and explained to you?<br><br>(This includes receiving the scan results in MyChart and the simple summary that was shared with you.) | ◯ Very satisfied<br>◯ Somewhat satisfied<br>◯ Neutral / No strong opinion<br>◯ Somewhat dissatisfied<br>◯ Very dissatisfied |
| When you reviewed this scan result and the provided simple summary, how did it affect your anxiety level? | ◯ I felt more anxious after reading the results and the simple summary<br>◯ I felt less anxious<br>◯ My anxiety level didn't really change<br>◯ I am not sure<br>◯ I prefer not to answer |
| How useful was the simple summary in helping you understand the medical information and scan results? | ◯ Extremely helpful<br>◯ Very helpful<br>◯ Somewhat helpful<br>◯ Slightly helpful<br>◯ Not helpful at all |

| | |
|---|---|
| Thinking about only the simple summary, how easy was it to understand the information presented in the simple summary? | ◯ Very easy to understand<br>◯ Somewhat easy to understand<br>◯ Neither easy nor difficult to understand<br>◯ Somewhat difficult to understand<br>◯ Very difficult to understand |
| Do you feel more prepared to discuss the results with your provider after reading this summary? | ◯ Yes<br>◯ No<br>◯ Unsure |
| Did this simple summary help answer your questions about the scan results?<br><br>(Select the option that best matches your experience.) | ◯ Yes. It answered all my questions, and I don't have any others.<br>◯ Yes. It answered some of my questions - I still have a few.<br>◯ Yes. It answered some of my questions - but I also have new questions now.<br>◯ No - it didn't answer the questions I had.<br>◯ I didn't have any questions to begin with. |
| After reviewing the simple summary, do you feel like you need to contact your provider with questions before your follow-up appointment with them? | ◯ Yes<br>◯ No |
| Did you notice any inaccuracies or other issues in the simple summary? | ◯ Yes<br>◯ No |
| Please describe the inaccuracies or other issues you noticed: | __________________________________ |
| Overall, how satisfied are you with the simple summary provided? | ◯ Very satisfied<br>◯ Somewhat satisfied<br>◯ Neutral / No strong opinion<br>◯ Somewhat dissatisfied<br>◯ Very dissatisfied |
| Suppose you continue to receive these simple summaries along with the radiology reports. Do you think you will need to contact your provider less frequently to ask questions or get clarifications on the scan results before your follow-up appointment with your cancer provider? | ◯ Yes<br>◯ No<br>◯ Unsure/Not applicable |
| Would you suggest that future imaging report communications include a simple summary like the one you reviewed? | ◯ Yes, definitely<br>◯ Yes, probably<br>◯ No, probably not<br>◯ No, definitely not |
| Based on the imaging summary you just reviewed, how would you describe your overall impression of the scan results?<br><br>(We ask this to better understand how different scan results may impact your feelings and stress levels.) | ◯ The scan results were good news<br>◯ The scan results were mixed<br>◯ The scan results were bad news<br>◯ I'm not sure how to interpret the results<br>◯ I prefer not to answer |
| What level of knowledge do you have of Artificial Intelligence (AI) in healthcare? | ◯ No knowledge<br>◯ Minimal knowledge<br>◯ Some knowledge<br>◯ A great deal of knowledge<br>◯ Expert-level knowledge |

| Question | Response |
| --- | --- |
| How comfortable are you with the use of AI in healthcare? | ◯ Very uncomfortable<br>◯ Uncomfortable<br>◯ Somewhat comfortable<br>◯ Comfortable<br>◯ Very comfortable |
| How appropriate does the use of AI to help prepare this scan summary feel to you? | ◯ Very appropriate<br>◯ Somewhat appropriate<br>◯ Unsure<br>◯ Somewhat inappropriate<br>◯ Very inappropriate |
| Did you see the AI disclosure statement in the simple scan summary? | ◯ Yes<br>◯ No<br>◯ Unsure |
| How do you feel about the way the use of AI was disclosed in the message you received with your scan summary?<br>(This is the disclaimer: This summary of your recent {Scan} is provided to help you understand the main findings in plain language. We used AI technology to help translate the radiology report into everyday terms, but the original image interpretation was performed by a radiologist. The AI did not read or interpret your scan images directly. I have carefully reviewed and edited this summary for accuracy before sharing it with you.) | ◯ The disclosure statement was clear and provided an appropriate level of detail.<br>◯ I understood the disclosure statement, but would have liked more details.<br>◯ The disclosure statement was confusing or unclear to me.<br>◯ I have concerns about how the use of AI was disclosed.<br>◯ Other (please explain below) |
| Please share your opinions on the AI disclosure statement and method. | __________________________________ |

### Claim your gift card

Please enter a valid email address if you would like to receive a $20 e-gift card. __________________________________

### Gift card distribution

Gift card was sent to the participant? __________________________________

Date when the gift card was sent. __________________________________

### Imaging Results Communication Survey

Thank you for your interest in this project. Before you get started, here's a brief overview of the project to help you understand what to expect from the survey.

What is this project about?

We are evaluating current practices and challenges around communicating scan results to patients. This research supports the rollout of a new AI-assisted Imaging Results Summary tool integrated within HealthLink. The tool uses a secure, UW Health-approved generative AI platform to createpatient-friendly summaries of radiology reports, aiming to improve scan result communication, improve patient understanding of the results and reduce their anxiety, and support provider workflows.
Your feedback will help us:

Understand how you currently communicate imaging results
Identify common workflow challenges
Refine the summary tool, designed workflow, and associated training materials

What will I be asked to do?

You will complete a brief survey (approx. 5 minutes) about your current imaging result communication practices and your perspectives on AI-assisted summaries.
No follow-up activities or additional tasks are required.

As a thank-you, participants will receive a $25 e-gift card (optional) for every completed survey.

Voluntary Participation and Confidentiality

Participation is voluntary, and you may skip any question or stop the survey at any time.If you choose to leave the study, your choice will not affect your employment or any services you receive. There will be no penalty to you.
Your responses will be confidential and used only for internal research and implementation purposes.
No identifying information will be published or shared outside of the study team.

Will my information be used in other research?

With appropriate confidentiality protections, we might use information that we collect during this study for other research or share it with other researchers without additional consent from you.

Questions?
If you have any questions, concerns, or complaints, or think that participating in the research has hurt you, please contact the PI:
□□ Toby Campbell, MD at 608-263-6002 or.

If you have concerns about your rights as a research participant or have complaints about the research study or study team, call the confidential research compliance line at 1-833-652-2506. UW Staff not part of the study team will work with you to address concerns and assist in resolving any complaints.

---

| Do you agree to participate in this survey? | ◯ Yes<br>◯ No |
|---|---|

### Imaging Results Communication Survey

| | |
|---|---|
| Approximately what percentage of your patients complete their scans on a different day before their scheduled follow-up visit with you? | ◯ 0-25%<br>◯ 26-50%<br>◯ 51-75%<br>◯ 76-100% |
| For patients who complete their scans before their office visit appointment, how often do you initiate communication about the results (e.g., phone, MyChart) prior to their follow-up visit? | ◯ Never<br>◯ Rarely<br>◯ Occasionally<br>◯ Frequently<br>◯ Very frequently |
| In the past month, how often did your patients contact you with questions or concerns about their scan results before their follow-up appointment? | ◯ Never<br>◯ Rarely<br>◯ Occasionally<br>◯ Frequently<br>◯ Very frequently |
| In the past month, how often have you initiated communication with your patients to discuss scan results before their upcoming appointment? | ◯ Never<br>◯ Rarely<br>◯ Occasionally<br>◯ Frequently<br>◯ Very frequently |
| In the past month, what is the impact of these scan-result-related communications on your clinical workload? | ◯ No impact<br>◯ Minimal impact<br>◯ Moderate impact<br>◯ Significant impact<br>◯ Overwhelming impact |
| If a patient-friendly draft summary was available for you to review and send to patients through MyChart, how often would you use it to communicate scan results? | ◯ Never<br>◯ Rarely<br>◯ Occasionally<br>◯ Frequently<br>◯ Always |

**What aspects of the patient-friendly summary tool would matter most to you in deciding whether to use it? (1st = the most important)**

| | 1st | 2nd | 3rd | 4th | 5th | 6th |
|---|---|---|---|---|---|---|
| Accuracy and clinical reliability of the content | ◯ | ◯ | ◯ | ◯ | ◯ | ◯ |
| Clarity and use of patient-friendly language | ◯ | ◯ | ◯ | ◯ | ◯ | ◯ |
| Seamless fit into my existing workflow | ◯ | ◯ | ◯ | ◯ | ◯ | ◯ |
| Time savings in communicating results | ◯ | ◯ | ◯ | ◯ | ◯ | ◯ |
| Positive impact on provider-patient communication | ◯ | ◯ | ◯ | ◯ | ◯ | ◯ |
| Ability to edit and personalize the summary | ◯ | ◯ | ◯ | ◯ | ◯ | ◯ |

| | |
|---|---|
| How useful do you think this tool could be for you and your patients? | ◯ Extremely helpful<br>◯ Very helpful<br>◯ Somewhat helpful<br>◯ Slightly helpful<br>◯ Not helpful at all |
| What level of knowledge do you have of Artificial Intelligence (AI) in healthcare? | ◯ No knowledge<br>◯ Minimal knowledge<br>◯ Some knowledge<br>◯ A great deal of knowledge<br>◯ Expert-level knowledge |
| How do you feel about the role of AI in medicine? | ◯ I don't trust it and prefer human expertise<br>◯ I am skeptical but open to learning more<br>◯ I am not sure. I need more information about how AI is used in medicine.<br>◯ I think AI has some potential, but it should be used cautiously<br>◯ I am very much in favor of AI being used more often in medicine. |
| How comfortable are you with automated tools drafting patient-facing communications (e.g., messages, summaries)? | ◯ Very uncomfortable<br>◯ Somewhat uncomfortable<br>◯ Neutral<br>◯ Somewhat comfortable<br>◯ Very comfortable |
| Do you currently use the Ambient AI-scribe (Abridge) for clinical documentation? | ◯ Yes, and I feel comfortable using AI for this purpose<br>◯ No, but I would feel comfortable trying one if/when available<br>◯ No, and I'm unsure about using AI tools for documentation<br>◯ No, and I prefer not to use AI for documentation<br>◯ No, I have used it in the past, but do not currently use it<br>◯ I'm not familiar with Ambient AI-scribe tools |
| What concerns, if any, do you have about using AI-generated content in clinical communication? (Select all that apply) | ☐ Accuracy of information<br>☐ Legal or liability risks<br>☐ Potential patient misunderstanding<br>☐ Integration into my workflow<br>☐ Ethical concerns<br>☐ I do not have concerns<br>☐ Other |
| Please describe your other concerns about using AI-generated content in clinical communication. | __________________________________ |
| Survey response status entered by the study team. | ◯ Invitation wasn't sent<br>◯ Provider didn't respond / Closed<br>◯ Partial response<br>◯ Completed |

### Provider AI-assisted Results Summary Feedback

| | |
|---|---|
| In the past month, how often did your patients contact you with questions or concerns about their scan results before their follow-up appointment? | ◯ Never<br>◯ Rarely<br>◯ Occasionally<br>◯ Frequently<br>◯ Very frequently |
| In the past month, how often have you initiated communication with your patients to discuss scan results before their upcoming appointment? | ◯ Never<br>◯ Rarely<br>◯ Occasionally<br>◯ Frequently<br>◯ Very frequently |
| In the past month, what has been the impact of the scan-result-related questions or messages from patients on your clinical workload? | ◯ No impact<br>◯ Minimal impact<br>◯ Moderate impact<br>◯ Significant impact<br>◯ Overwhelming impact |
| How many times have you sent one of these AI-drafted imaging report summaries to your patients so far? | ◯ 0<br>◯ 1-5<br>◯ 6-10<br>◯ >10 |
| How would you rate the overall quality of the summaries you reviewed? | ◯ Very good<br>◯ Good<br>◯ Fair<br>◯ Poor<br>◯ Very poor |
| Did you notice any significant issues or inaccuracies in the summaries? | ◯ No<br>◯ Yes (please describe) |
| Please describe the issues or inaccuracies you noticed in the summaries. | __________________________________ |
| How much do you trust the clinical accuracy of the AI-drafted summaries before editing? | ◯ Completely<br>◯ Mostly<br>◯ Somewhat<br>◯ Not at all |
| What challenges did you face when using the summary tool? (Select all that apply) | ☐ Not aware of this tool<br>☐ Workflow issues (doesn't fit into my clinical workflows)<br>☐ Trust in content<br>☐ Time constraints<br>☐ Other |
| Please describe the other challenges you experienced. | __________________________________ |
| How easy or difficult was it to use the designed workflow to send summaries to patients? | ◯ Very easy<br>◯ Somewhat easy<br>◯ Somewhat difficult<br>◯ Very difficult |

| | |
|---|---|
| How satisfied are you with the current workflow for reviewing and sending these summaries to patients? | ◯ Very satisfied<br>◯ Somewhat satisfied<br>◯ Somewhat unsatisfied<br>◯ Very unsatisfied |
| What do you like about the workflow, and what could be improved? | __________________________________ |
| What is the impact of these AI-drafted summaries on your imaging results communication with your patients? | ◯ Positive impact; they improved my communication<br>◯ Slight positive impact; a small but helpful improvement<br>◯ No impact; my communication remains unchanged<br>◯ Slight negative impact; they made communication slightly more difficult<br>◯ Negative impact; they interfered with my communication |
| Did you use or reference the summaries during in-person or virtual visits? | ◯ Yes - often<br>◯ Yes - occasionally<br>◯ No |
| Have you received any feedback from patients about the summaries? | ◯ No<br>◯ Yes, I have received mostly positive feedback<br>◯ Yes, I have received mixed feedback<br>◯ Yes, I have received mostly negative feedback |
| Please describe the feedback you received from patients. | __________________________________ |
| To what extent do you agree with the following statement: "The summaries helped my patients better understand their imaging results." | ◯ Strongly agree<br>◯ Agree<br>◯ Neutral<br>◯ Disagree<br>◯ Strongly disagree |
| What is the impact of these AI-drafted summaries on your scan result-related communication workload? | ◯ No impact<br>◯ Minimal impact<br>◯ Moderate impact<br>◯ Significant impact |
| Please describe how the summary tool changed your documentation or communication time. | __________________________________ |
| Would you recommend continuing to use the summary tool for communicating scan results? | ◯ Yes<br>◯ No<br>◯ Not sure |
| Please share your comments on this recommendation. | __________________________________ |
| How likely are you to continue using the AI-assisted summary tool to communicate scan results in your future practice? | ◯ Very unlikely<br>◯ Unlikely<br>◯ Not sure<br>◯ Likely<br>◯ Very likely |

| | |
|---|---|
| What level of knowledge do you have of Artificial Intelligence (AI) in healthcare? | ◯ No knowledge<br>◯ Minimal knowledge<br>◯ Some knowledge<br>◯ A great deal of knowledge<br>◯ Expert-level knowledge |
| How do you feel about the role of AI in medicine? | ◯ I don't trust it and prefer human expertise<br>◯ I am skeptical but open to learning more<br>◯ I am not sure. I need more information about how AI is used in medicine.<br>◯ I think AI has some potential, but it should be used cautiously<br>◯ I am very much in favor of AI being used more often in medicine. |
| How has your attitude toward using AI-generated content in patient care changed after this experience? | ◯ More positive<br>◯ No change<br>◯ More negative |
| If you identified a change in your attitude, what contributed most to this change? | ________________________________ |
| Survey response status entered by the study team | ◯ Invitation wasn't sent<br>◯ Provider didn't respond / Closed<br>◯ Partial response<br>◯ Completed |

### Claim your gift card

Please enter a valid email address if you would like to receive a $25 e-gift card. ______________________________

### Manual advance

Survey sent date entered by staff

__________________________________

Advance to the next event.

◯ Yes
◯ No
